# PARIS (Pneumonia: Acute Respiratory Infection ± Sepsis): a prospective single-centre observational cohort study of hospitalised patients with pneumonia

**DOI:** 10.64898/2026.07.15.26357955

**Authors:** Syed Tahir Nasser, Charlie Piercy, Agnieszka Falinska, Fernando Martinez-Estrada, Denise M. O’Sullivan, Alison Devonshire, Jim Huggett, Ben Creagh-Brown

## Abstract

**Introduction:** Hospitalised community-acquired pneumonia (CAP) is heterogeneous in aetiology, severity, and outcome. Phenotyping and endotyping approaches offer potential to stratify patients biologically and guide targeted therapy, but require well-characterised cohorts with linked biosamples. We describe the PARIS (Pneumonia: Acute Respiratory Infection ± Sepsis) study: a prospective observational cohort of hospitalised patients with pneumonia, designed to characterise functional outcomes and to provide a biobank for translational immunological research.

**Methods:** Adults admitted with CAP to a single NHS district general hospital were enrolled within 24 hours of admission between December 2020 and March 2022. Clinical, functional, and physiological data were collected at enrolment, hospital discharge, and 6–8 week follow-up. Serial blood samples were collected for flow cytometry, transcriptomics, pathogen DNA detection, and plasma biobanking.

**Results:** Forty-seven patients were enrolled (15 without and 32 with sepsis [SOFA ≥2] at enrolment); 87% met sepsis criteria by 24 hours post enrolment. Most patients (30/47, 64%) were managed as COVID-19, microbiologically confirmed in 27. Mean age was 57 years (SD 16), 70% were male, and baseline comorbidity burden was low. Severity was moderate (median NEWS2 4 at enrolment, rising to 6 by 24 hours post enrolment; p<0.001). Mortality was 4/47 (8.5%), with 44/47 (94%) alive at hospital discharge. Median length of stay was 8 days (IQR 5.5–11). Translational samples were collected from the majority: fresh flow cytometry (44/47, 94%), transcriptomics from the sepsis subgroup (31/32, 97%), pathogen DNA sampling (35 samples received across study timepoints; see Table 5), and stored plasma (29/47, 62%). The primary outcome of functional decline (Barthel score decrease ≥1.85) occurred in only 1/29 patients with paired assessments (3.4%). Persistent CRP elevation (>3 mg/L) at 6–8 week follow-up was present in 16/31 (52%) survivors with available data.

**Conclusions:** The PARIS cohort provides a well-characterised clinical platform and linked biobank to support translational studies of pneumonia and sepsis. The low rate of functional decline reflects the younger, lower-comorbidity, COVID-predominant population recruited. Primary protocol endpoints were not achieved owing to pandemic-related disruption. Data and samples underpin a programme of linked translational studies.

## Introduction

Community-acquired pneumonia (CAP) was among the leading causes of infectious disease mortality in western countries prior to the COVID-19 pandemic and remains a major burden on healthcare resources.(1) It is one of the most frequent precipitants of sepsis and acute respiratory distress syndrome (ARDS), and hospitalisation for CAP is increasingly common.(1)

Despite sharing a common anatomical diagnosis, pneumonia is highly heterogeneous in aetiology, severity, and host response. It is increasingly recognised that this clinical heterogeneity reflects underlying biological variation: patients can be grouped into clinically meaningful **phenotypes** based on observable characteristics, and into **endotypes** based on the distinct biological mechanisms driving their illness. Analysis of transcriptomic data from major sepsis cohorts, many of whom have CAP as the original diagnosis, has identified biologically and prognostically distinct immune endotypes, with differing predicted responses to immunomodulatory treatment. The Genomic Advances in Sepsis (GAinS) consortium identified two such endotypes in patients with CAP (termed SRS1 and SRS2),(2) while the Molecular Diagnosis and Risk Stratification of Sepsis (MARS) consortium described four endotypes in a broader all-cause sepsis population.(3)(4) Analogous subphenotypes have been characterised in ARDS using clinical and biomarker data.(5) Advancing this field requires well-characterised cohorts with linked biosamples that enable retrospective and prospective endotyping analyses.

Beyond the acute illness, short- and long-term morbidity are substantially increased following hospitalisation for CAP. Many patients leave hospital with ongoing subclinical inflammation, which is associated with subsequent mortality.(6) Persistent systemic inflammation following critical illness is associated with impaired recovery of physical function.(7) Cross-sectional studies have consistently found associations between elevated inflammatory markers and frailty.(8) A significant proportion of patients still experience symptoms up to 90 days following CAP, and survivors of sepsis frequently cannot resume previous roles and activities for months after discharge.(9)(10) Understanding the pathophysiological mechanisms linking acute CAP with its long-term sequelae has been highlighted as a research priority.(11)

There is relative scarcity of published data on recovery of physical function and persistence of systemic inflammation following hospitalisation with CAP, or on the routine provision of physical rehabilitation and nutritional support. We designed the PARIS (Pneumonia: Acute Respiratory Infection ± Sepsis) study as a prospective observational cohort with two principal aims: to characterise functional outcomes and the persistence of systemic inflammation following hospitalisation for CAP; and to collect serial biosamples to support a programme of translational immunological studies including immune phenotyping and endotyping. This paper describes the clinical cohort and biobank, serving as a methods reference for linked translational studies.

## Methods

### Design and setting

We conducted a single-centre prospective observational cohort study (PARIS) recruiting between 15th December 2020 and 25th March 2022 at Royal Surrey County Hospital (RSCH), a medium-sized NHS district general hospital in Guildford, Surrey, UK. The study was part of the SEPTIMET programme (‘Metrology for clinical implementation of sepsis biomarkers’), funded under the European Metrology Programme for Innovation and Research (EMPIR), which aimed to standardise and improve the accuracy of clinical measurements for sepsis and lower respiratory tract infection biomarkers. Biomarker metrological aspects are not reported within this manuscript.

### Protocol modifications due to COVID-19

The study was designed prior to the COVID-19 pandemic and had originally been intended to run at six UK centres with a target enrolment of approximately 500 patients over 18 months. Pandemic-related disruption necessitated a single-centre design with a substantially reduced sample size. SARS-CoV-2 was retained as an eligible aetiology to maintain feasible recruitment.

### Study objectives

**Primary objectives:** (i) Immunodiagnostic: to assess metrological characteristics of procalcitonin (PCT) as a sepsis biomarker; (ii) Functional: to determine the proportion of patients with increased functional dependency (Barthel index decrease ≥1.85) at 6–8 week follow-up.

**Secondary objectives:** persistence of systemic inflammation (CRP >3 mg/L and >10 mg/L at follow-up); association between persistent inflammation and functional decline; changes in handgrip strength, CAP symptom score (CAPsym), Duke Activity Status Index (DASI), and Clinical Frailty Scale; nutritional risk and support; receipt of physical assistance during admission; determinants of adverse outcomes; and long-term mortality via national data linkage. The PCT primary objective was not achieved owing to pandemic-related disruption to the international SEPTIMET collaboration.

### Eligibility criteria

We recruited adults (≥18 years) admitted to hospital within 24 hours of symptom onset with a predicted stay of at least 48 hours who fulfilled a pre-specified definition of CAP based on a minor adaptation of the definition used in the CDC Etiology of Pneumonia in the Community (EPIC) study,(1) compatible with national guidelines.(12) The CAP definition required all of the following: (i) evidence of acute infection (fever, chills, leucocytosis, leukopenia, or new altered mental status); (ii) evidence of acute respiratory illness (new cough, sputum production, chest pain, dyspnoea, tachypnoea, abnormal lung examination, or respiratory failure); and (iii) radiographic or CT evidence of pneumonia.

Exclusion criteria were: immunocompromised (HIV, chemotherapy, or long-term immunomodulating medications including corticosteroids); suspicion of active tuberculosis; currently incarcerated; inadequate English language proficiency; palliative or community DNACPR in place; significant anaemia (Hb <80 g/L); and pregnancy.

Patients were subclassified at enrolment into those with and without sepsis (SOFA score ≥2 at enrolment),(13) targeting 30 patients in each group. As pre-specified in the protocol, the two groups are combined for all analyses in this paper.

### Consent

Written informed consent was granted directly where possible. For patients lacking capacity at enrolment, agreement from a personal consultee was sought, followed by retrospective consent on return of capacity. Where patients did not regain capacity, previously collected data was retained inline with the personal consultee agreement, no additional data was collected.

### Schedule of assessments

**Enrolment (within 24 hours of admission):** assessment of handgrip strength, DASI, Barthel index, CFS, and CAPsym(14) — a validated 18-item patient-reported measure of symptom burden in CAP, primarily used in antibiotic and anti-inflammatory trials;(15)(16) its serial use in observational cohorts is relatively uncommon.(17) CRP was measured as part of routine clinical care.

**At hospital discharge or day 7 (whichever sooner):** handgrip strength and CAPsym. CRP measured if not already obtained clinically. ICU Mobility Score(18) recorded daily during admission.

**At 6–8 week follow-up (V3):** handgrip strength, DASI, Barthel index, CFS, CAPsym, CRP, and chest radiograph as per national guidelines.

Receipt of mobilisation assistance and nutritional support was recorded daily during admission. These data were collected to characterise current practice and to inform the design of future interventional trials in this setting; this is an aspect of CAP management that is rarely captured systematically in observational studies. Malnutrition risk was assessed using the Malnutrition Universal Screening Tool (MUST).

### Blood sample collection

At enrolment, blood samples were collected from all participants into EDTA and serum separator tubes for immunophenotyping (fresh flow cytometry), biomarker assessment, and plasma biobanking. Participants meeting sepsis criteria at enrolment underwent additional sampling into PAXgene Blood RNA tubes (PreAnalytiX) for leucocyte RNA transcriptomic analysis, together with samples for pathogen DNA detection and procalcitonin (PCT) measurement, with PCT analysed in the Royal Surrey NHS pathology laboratory. Samples were processed within the Royal Surrey Research Laboratory according to study-specific standard operating procedures and stored at −80°C pending analysis. Serial blood sampling was performed in the sepsis subgroup, including repeat collection at hospital discharge. Planned paired reference laboratory PCT measurements (Laboratoire National de Métrologie et d’Essais, Paris) were not undertaken owing to pandemic-related disruption to international research activities. Detailed laboratory methods and analyses are reported in linked publications.

### Statistical analysis

The original study design was powered to detect a 20% absolute difference in the proportion of patients with Barthel index decline ≥1.85, assuming 25% in the CAP group and 5% in a planned healthy control group, at 80% power and 5% significance (two-tailed), requiring approximately 70 patients per group. The healthy control group was not recruited and this sample size was not achieved, owing to pandemic-related disruption. This paper therefore presents descriptive statistics for the cohort as a whole. Continuous variables are reported as mean (SD) or median (IQR) as appropriate; categorical variables as counts and percentages. Within-patient comparisons across timepoints used the Wilcoxon matched-pairs signed-rank test. The primary functional outcome — proportion with Barthel decrease ≥1.85 at follow-up — is reported with a

95% confidence interval (exact binomial). Missing data are reported without imputation. Analyses were performed in Python (version 3.12; pandas, scipy, matplotlib).

### Ethics and registration

Ethical approval: South Central – Hampshire A Research Ethics Committee, REC reference 20/SC/0006, 16 November 2020. IRAS project ID: 269427. Sponsored by Royal Surrey NHS Foundation Trust; listed on the NIHR portfolio. The study was not registered on a prospective clinical trial registry, which is acknowledged as a limitation.

## Results

### Recruitment and cohort overview

Forty-seven patients were enrolled between 15th December 2020 and 25th March 2022. Fifteen did not fulfil sepsis criteria (SOFA ≥2) at enrolment and 32 did. The proportion meeting sepsis criteria rose to 87% (41/47) by 24 hours post enrolment (Figure 1). Per protocol, the two enrolment groups are combined for all analyses.

**Figure 1.**
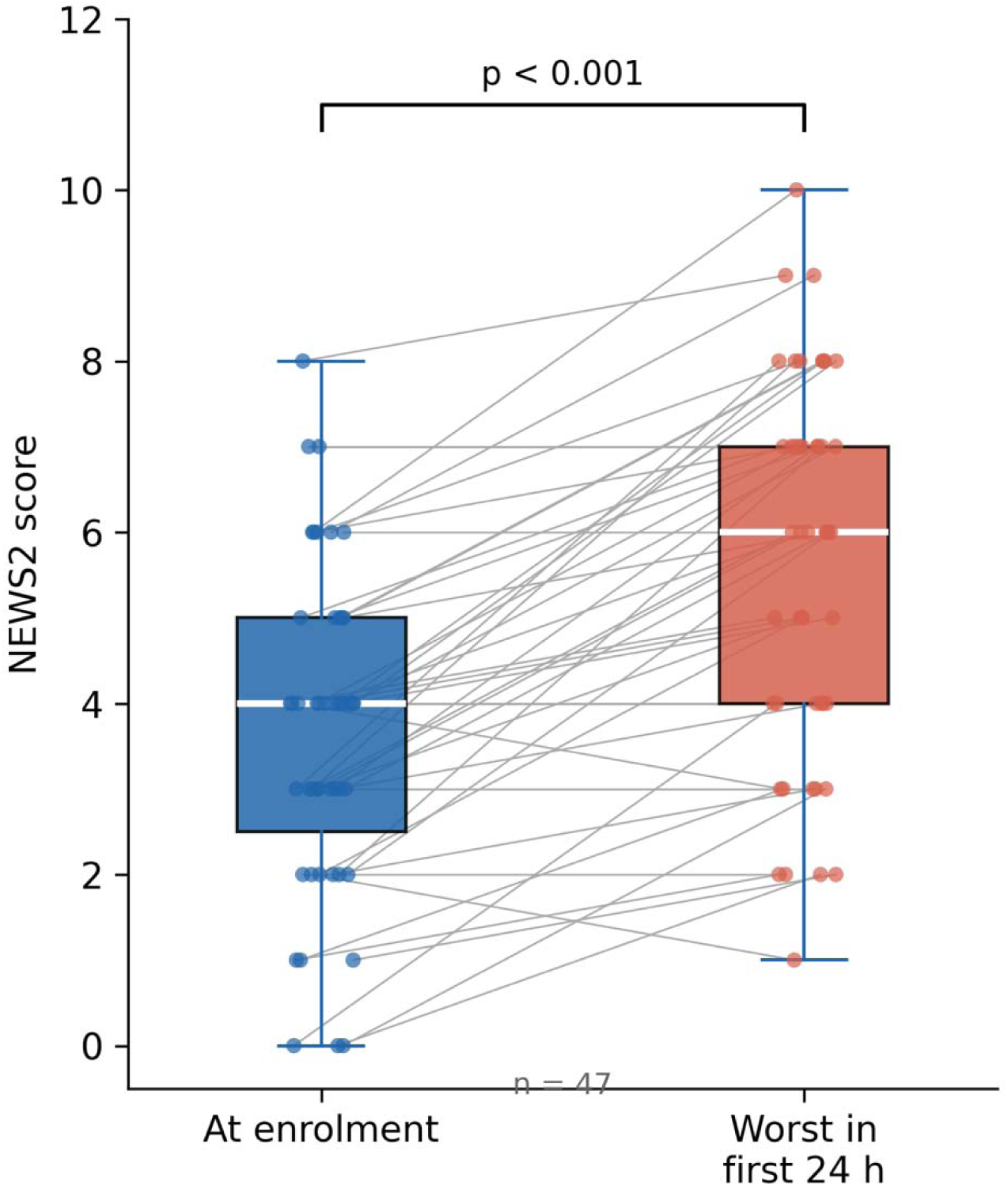
Change in NEWS2 score between enrolment and the highest value recorded in the 24 hours post enrolment. Individual patient values are shown with connecting lines (grey); boxes show median and IQR (blue = enrolment; red = 24 h post enrolment). NEWS2: National Early Warning Score 2. Wilcoxon matched-pairs signed-rank test, p<0.001 (n=47).

### Patient characteristics

Demographic and baseline characteristics are shown in Table 1. The cohort was relatively young (mean age 57 years, SD 16) with low comorbidity burden; all patients lived in their own home prior to admission. Most were functionally independent at baseline: median Barthel index 20 (fully independent) and median CFS 2. A minority had received antibiotics in the 7 days prior to admission (9/47, 19%) and a small number were taking inhaled corticosteroids (4/47, 9%).

**Table 1:**
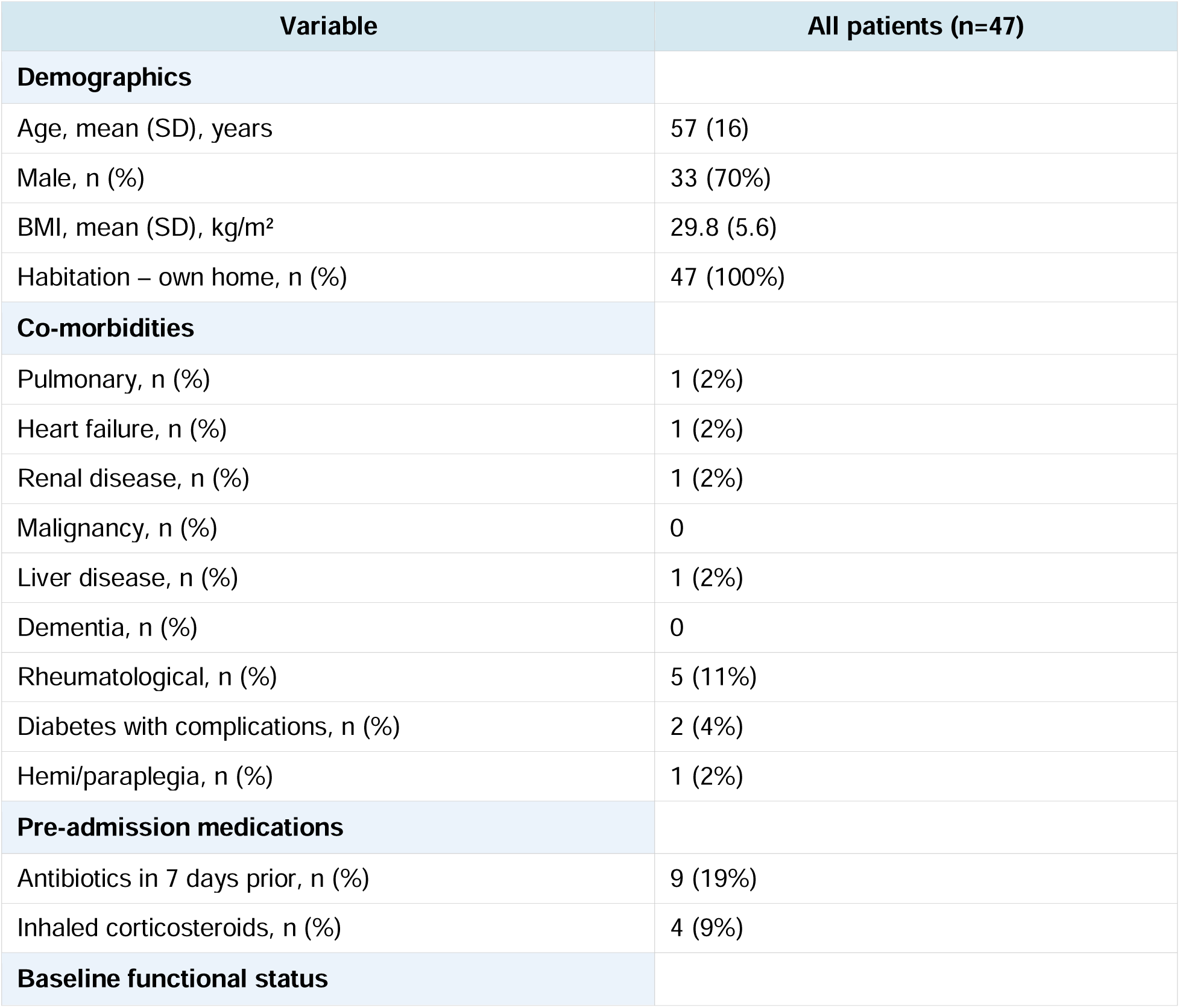
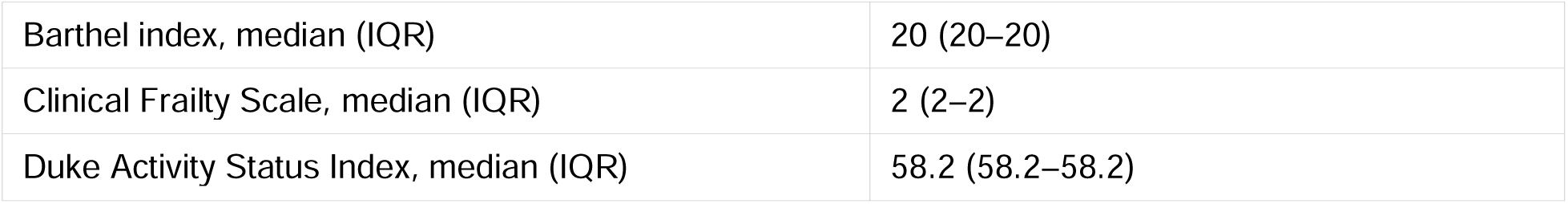
Demographic and baseline characteristics (n=47).

| Variable | All patients (n=47) |
| --- | --- |
| **Demographics** |  |
| Age, mean (SD), years | 57 (16) |
| Male, n (%) | 33 (70%) |
| BMI, mean (SD), kg/m<sup>2</sup> | 29.8 (5.6) |
| Habitation – own home, n (%) | 47 (100%) |
| **Co-morbidities** |  |
| Pulmonary, n (%) | 1 (2%) |
| Heart failure, n (%) | 1 (2%) |
| Renal disease, n (%) | 1 (2%) |
| Malignancy, n (%) | 0 |
| Liver disease, n (%) | 1 (2%) |
| Dementia, n (%) | 0 |
| Rheumatological, n (%) | 5 (11%) |
| Diabetes with complications, n (%) | 2 (4%) |
| Hemi/paraplegia, n (%) | 1 (2%) |
| **Pre-admission medications** |  |
| Antibiotics in 7 days prior, n (%) | 9 (19%) |
| Inhaled corticosteroids, n (%) | 4 (9%) |
| **Baseline functional status** |  |
| Barthel index, median (IQR) | 20 (20–20) |
| Clinical Frailty Scale, median (IQR) | 2 (2–2) |
| Duke Activity Status Index, median (IQR) | 58.2 (58.2–58.2) |

### Severity and clinical course

Severity of illness at enrolment and treatments received are summarised in Table 2. Physiological disturbance was moderate at enrolment (median NEWS2 4, IQR 3–5). The highest NEWS2 in the 24 hours post enrolment was significantly greater (median 6, IQR 4–7; Wilcoxon p<0.001; Figure 1). Most patients required supplemental oxygen via face mask or nasal cannulae (33/43 with data, 77%); 9 (21%) required CPAP or non-invasive ventilation and 1 (2%) required high-flow nasal oxygen. No patient required invasive mechanical ventilation. Six patients were admitted to critical care, with a median ICU length of stay of 14.5 days (IQR 6– 24). Three patients died in hospital within 30 days of enrolment; one further patient died after discharge between 30 days and 10 weeks post-enrolment, giving a total of 44/47 (94%) alive at hospital discharge.

**Table 2:**
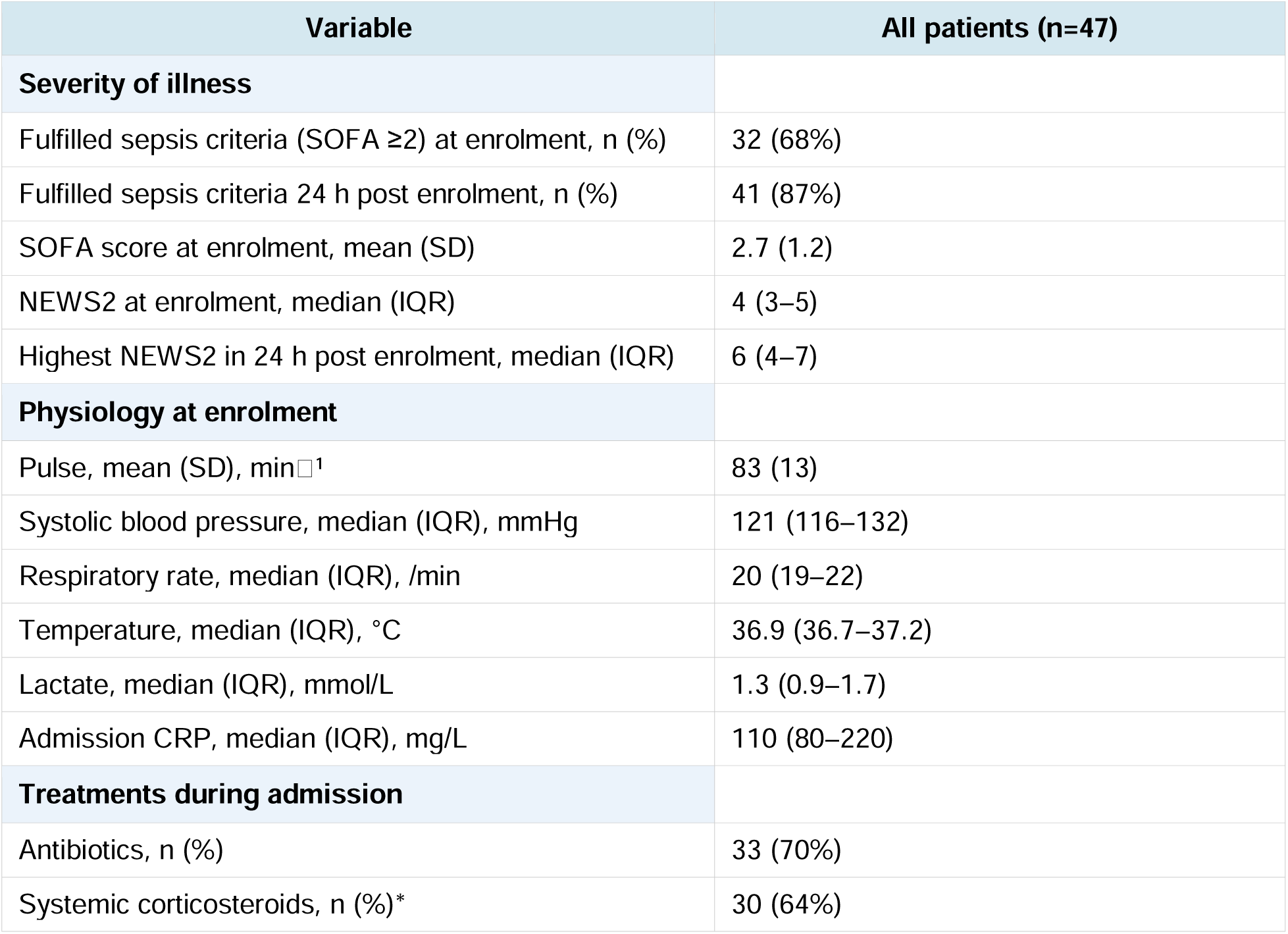
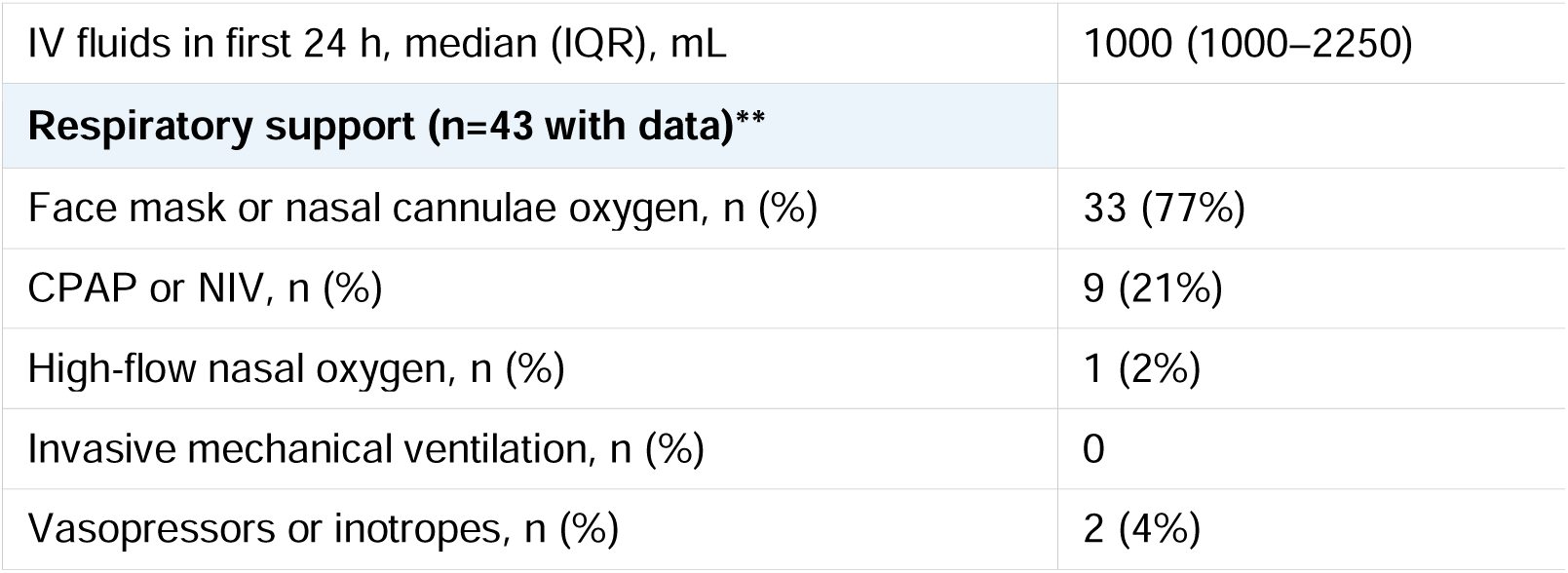
Severity of acute illness and treatments (n=47). *High proportion reflects COVID-19 management guidelines operative throughout the recruitment period (see Discussion). **4 patients had missing respiratory support data.

| Variable | All patients (n=47) |
| --- | --- |
| **Severity of illness** |  |
| Fulfilled sepsis criteria (SOFA $\geq 2$) at enrolment, n (%) | 32 (68%) |
| Fulfilled sepsis criteria 24 h post enrolment, n (%) | 41 (87%) |
| SOFA score at enrolment, mean (SD) | 2.7 (1.2) |
| NEWS2 at enrolment, median (IQR) | 4 (3–5) |
| Highest NEWS2 in 24 h post enrolment, median (IQR) | 6 (4–7) |
| **Physiology at enrolment** |  |
| Pulse, mean (SD), min<sup>-1</sup> | 83 (13) |
| Systolic blood pressure, median (IQR), mmHg | 121 (116–132) |
| Respiratory rate, median (IQR), /min | 20 (19–22) |
| Temperature, median (IQR), °C | 36.9 (36.7–37.2) |
| Lactate, median (IQR), mmol/L | 1.3 (0.9–1.7) |
| Admission CRP, median (IQR), mg/L | 110 (80–220) |
| **Treatments during admission** |  |
| Antibiotics, n (%) | 33 (70%) |
| Systemic corticosteroids, n (%)* | 30 (64%) |
| IV fluids in first 24 h, median (IQR), mL | 1000 (1000–2250) |
| **Respiratory support (n=43 with data)\*\*** |  |
| Face mask or nasal cannulae oxygen, n (%) | 33 (77%) |
| CPAP or NIV, n (%) | 9 (21%) |
| High-flow nasal oxygen, n (%) | 1 (2%) |
| Invasive mechanical ventilation, n (%) | 0 |
| Vasopressors or inotropes, n (%) | 2 (4%) |

### Inflammation

Admission CRP was elevated in all patients (median 110 mg/L, IQR 80–220). CRP fell substantially by hospital discharge (median 28 mg/L, IQR 14.5–41.5; n=31) and further at 6–8 week follow-up (median 8.8 mg/L, IQR 7.6–12.5; n=16; Figure 2A). Of 31 survivors with a follow-up CRP result, 16 (52%) had persistently elevated CRP (>3 mg/L), including 6 (19%) with values >10 mg/L. Among the 15 patients with both a follow-up chest radiograph and a follow-up CRP value, there was no significant difference in CRP between those with and without persistent radiological opacification (median 8.9 vs 8.6 mg/L; Mann-Whitney U, p=0.40); the small number limits any inference from this comparison. In an exploratory comparison, admission CRP was significantly higher in patients not managed as COVID-19 (median 220 mg/L, IQR 110–325) than in those managed as COVID-19 (median 101 mg/L, IQR 66–157; Mann-Whitney U, p=0.006).

**Figure 2.**
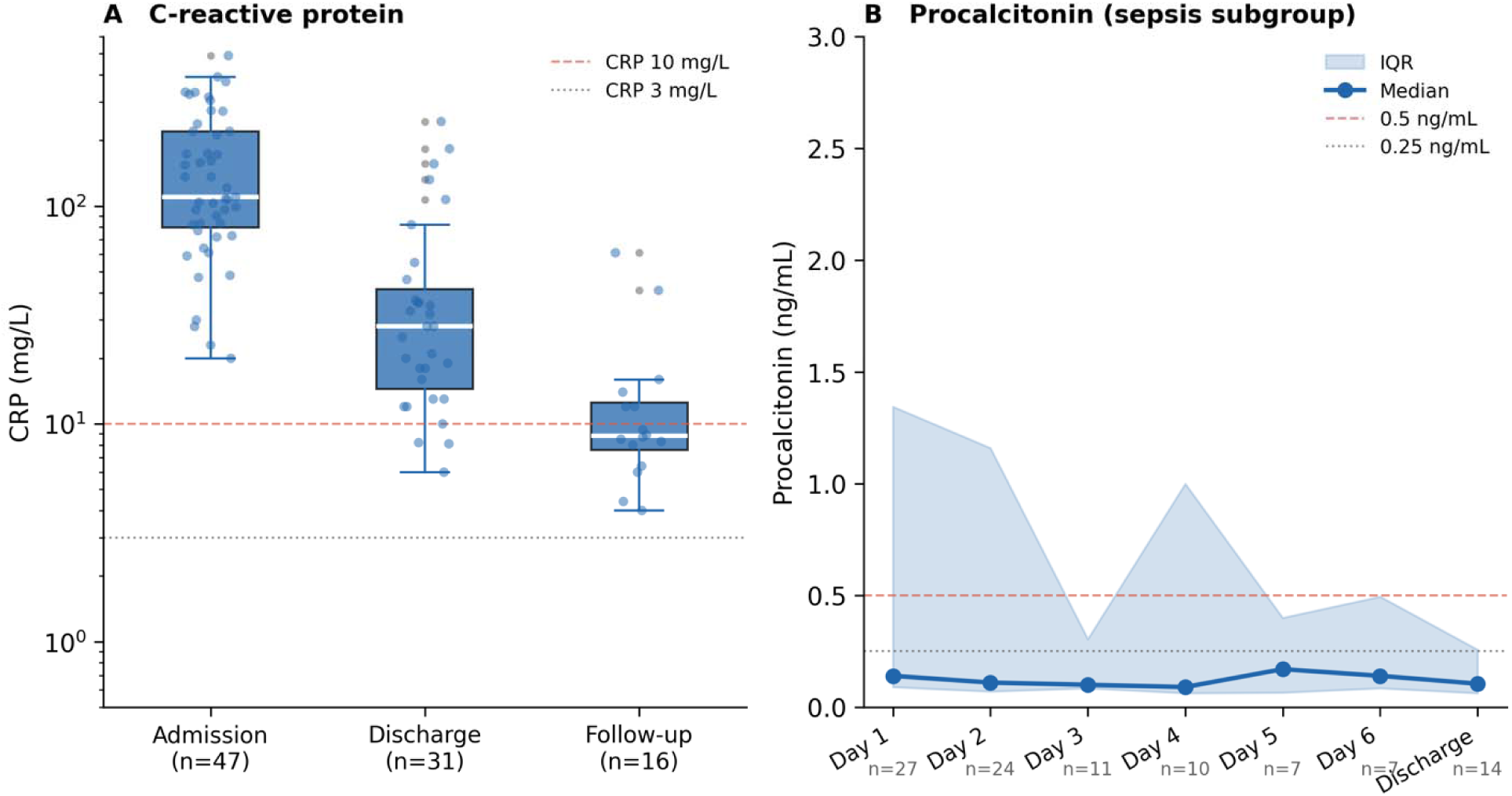
A) C-reactive protein (CRP) at admission, hospital discharge, and 6–8 week follow-up. Individual data points overlaid on box plots (median and IQR). Dashed red line: 10 mg/L; dashed grey line: 3 mg/L. Sample sizes: admission n=47, discharge n=31, follow-up n=16. B) Procalcitonin (PCT) measured in the NHS clinical laboratory in patients fulfilling sepsis criteria at enrolment. Line: median values; shaded region: IQR. Dashed red line: 0.5 ng/mL (sepsis threshold); dashed grey line: 0.25 ng/mL. Sample sizes per timepoint shown below the x-axis.

Procalcitonin (PCT) was measured in the NHS clinical laboratory in patients meeting sepsis criteria at enrolment (n=27 on day 1). Admission values were low (median 0.14 ng/mL, IQR 0.09–1.34) and showed rapid resolution over the first days of admission, with values remaining below 0.5 ng/mL in the majority throughout (Figure 2B).

### Resolution of symptom burden

CAPsym(14) scores were markedly elevated at enrolment (median 38, IQR 29–47; n=42), broadly consistent with values reported in recent CAP trials.(15)(16) Scores fell substantially by hospital discharge (median 15, IQR 0–25; n=47) but did not improve further at follow-up (median 18, IQR 11–29; n=31; Figure 3).

**Figure 3.**
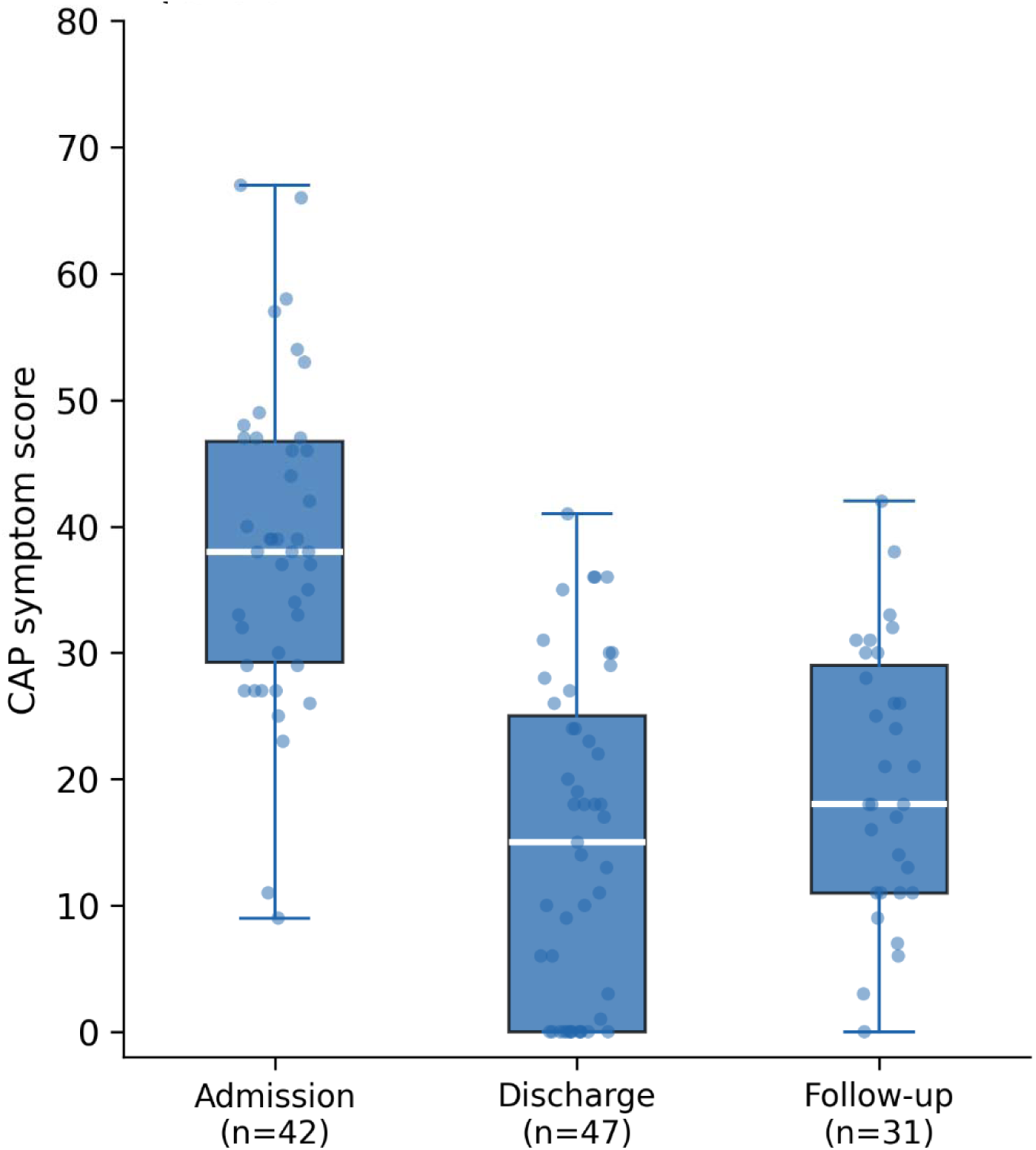
CAP symptom score (CAPsym) at admission, hospital discharge, and 6–8 week follow-up. Individual data points overlaid on box plots (median and IQR). Higher scores indicate greater symptom burden (18-item scale, each item scored 020135; maximum 90). Sample sizes: admission n=42, discharge n=47, follow-up n=31.

### Functional measures

There were high rates of missing data for functional measures at follow-up; four patients died before the planned V3 assessment.

The DASI showed a lower median at follow-up (42.7, n=31) compared with enrolment (58.2, n=39; Wilcoxon p=0.21 in 25 paired patients). The proportion achieving the maximum DASI score (58.2) decreased from 51% (20/39) at enrolment to 35% (11/31) at follow-up.

The primary functional outcome — proportion with Barthel index decrease ≥1.85 between enrolment and follow-up — occurred in 1 of 29 patients with paired assessments (3.4%; 95% CI 0.1–10.3%). Functional independence was high throughout: 87% were fully independent (Barthel 20) at enrolment (39/45) and 87% at follow-up (27/31), in those with available data.

The CFS showed a median of 2 at both timepoints. The proportion scoring CFS 1 (very fit) decreased from 13% (6/46) at enrolment to 6% (2/31) at follow-up. Return-to-work data were too incomplete for meaningful analysis.

Handgrip strength improved from enrolment (median 34 kg, IQR 21–42; n=46) to discharge (median 37 kg, IQR 30–46; Wilcoxon p=0.002; n=29 paired; Figure 4). ICU Mobility Scores were available in most patients: 54% required some degree of assistance (score <10) at enrolment, falling to 24% at discharge. At follow-up, 55% (18/33) of those assessed still required some degree of assistance.

**Figure 4.**
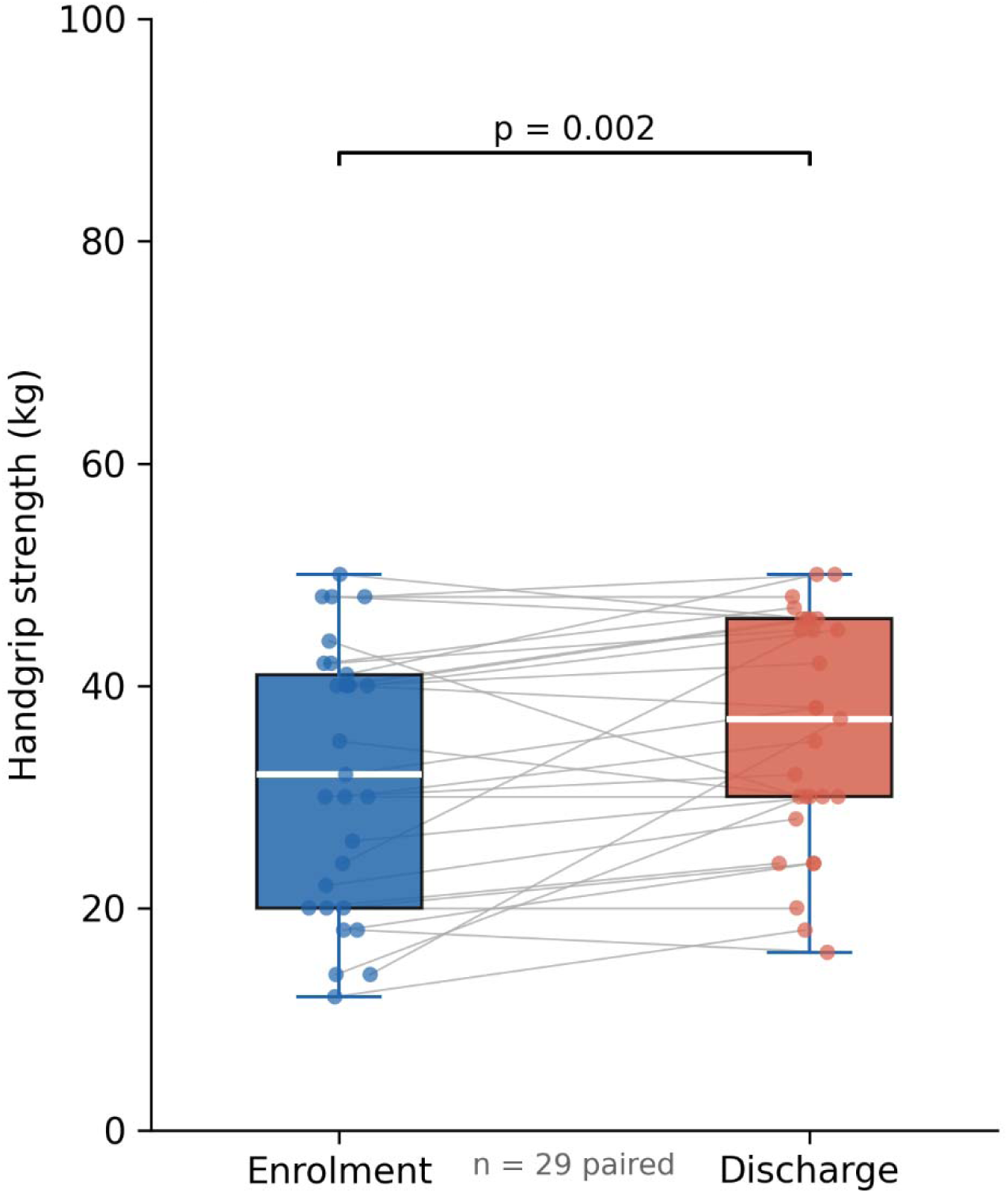
Handgrip strength (dominant hand) at enrolment and hospital discharge. Individual patient values shown with connecting lines (grey). Blue = enrolment; red = discharge. Boxes show median and IQR. Wilcoxon matched-pairs signed-rank test, p=0.002 (n=29 paired).

### Microbiology and clinical outcomes

Most patients (30/47, 64%) were managed as COVID-19, microbiologically confirmed in 27. *Streptococcus pneumoniae* was confirmed from blood cultures in 3 patients, and 1 had an alternative confirmed organism. No causative agent was identified in 16 patients (34%). Clinical outcomes are summarised in Table 3.

**Table 3:** Causative organisms and clinical outcomes (n=47). †One further patient died after discharge between 30 days and 10 weeks post-enrolment. CXR denominator: patients who survived and had a CXR performed (n=39); 4 patients died, 3 had no CXR, 1 missing.

| Variable | All patients (n=47) |
| --- | --- |
| **Causative organism** |  |
| Managed as COVID-19, n (%) | 30 (64%) |
| Microbiologically confirmed COVID-19, n | 27 |
| *Streptococcus pneumoniae* (blood culture), n | 3 |
| Other confirmed organism, n | 1 |
| No causative agent confirmed, n (%) | 16 (34%) |
| **Clinical outcomes** |  |
| Admission to critical care, n (%) | 6 (13%) |
| ICU length of stay (ICU patients), median (IQR), days | 14.5 (6–24) |
| Total hospital length of stay, median (IQR), days | 8 (5.5–11) |
| Alive at hospital discharge†, n (%) | 44 (94%) |
| Total mortality within 10 weeks of enrolment, n (%) | 4 (8.5%) |
| Death within 30 days of enrolment (in-hospital), n | 3 |
| Death after 30 days but within 10 weeks, n | 1 |
| **Complications during admission** |  |
| Delirium, n (%) | 4 (9%) |
| Atrial fibrillation, n (%) | 4 (9%) |
| **Radiological follow-up** |  |
| Persistent opacification on follow-up CXR, n/N with CXR (%) | 17/39 (44%) |

### Nutritional assessment and support

Malnutrition risk screening (MUST) was completed in 45 of 47 patients (96%); the majority were at low risk (MUST 0: 36/45, 80%). Dietitian review or nutritional supplementation was documented in 91% of patients; only 4 (9%) had no record of either. Table 4 summarises nutritional data.

**Table 4:** Nutritional risk screening and support (n=47). 2020Percentage of patients with MUST completed (n=45).

| Variable | All patients (n=47) |
| --- | --- |
| **Malnutrition Universal Screening Tool (MUST)** |  |
| MUST not completed, n (%) | 2 (4%) |
| MUST = 0 (low risk), n (%)2020 | 36/45 (80%) |
| MUST = 1 (medium risk), n (%) | 5 (11%) |
| MUST ≥ 2 (high risk), n (%) | 4 (9%) |
| **Nutritional support** |  |
| Reviewed by dietitian within first 24 h, n (%) | 24 (51%) |
| No record of dietitian review or supplementation, n (%) | 4 (9%) |

**Table 5:** Translational biosample collection. †Sepsis subgroup (n=32) only, per protocol. ‡Clinical NHS laboratory; reference laboratory paired measurements were not undertaken. §35 pathogen DNA samples were received by the analysing laboratory: 13 at day 2, 11 at enrolment (within 24 h), 11 at discharge or day 7 (whichever sooner).

| Sample type | Timepoint | n collected | % of eligible |
| --- | --- | --- | --- |
| Fresh flow cytometry | Enrolment | 44 | 94% (44/47) |
| Transcriptomics (leucocyte RNA)† | Enrolment | 31 | 97% (31/32) |
| Pathogen DNA (samples received)§ | — | 35 | — |
| Plasma (stored) | Enrolment | 29 | 62% (29/47) |
| Plasma (stored) | Discharge | 22 | 47% (22/47) |
| Procalcitonin (clinical laboratory)†‡ | Enrolment (day 1) | 27 | 84% (27/32) |
| Procalcitonin (clinical laboratory)†‡ | Days 2–6 and discharge | 7–24 | See Figure 2B |

### Translational biosample collection

Comprehensive blood sampling was achieved in the majority of eligible patients (Table 5). These samples, together with the clinical characterisation above, provide the platform for translational analyses detailed in linked publications.

## Discussion

The PARIS cohort enrolled 47 patients hospitalised with pneumonia during endemic SARS-CoV-2 transmission — substantially fewer than planned and at a single centre rather than the intended six. Despite these modifications, comprehensive translational sample collection was achieved in the majority of patients. The primary functional endpoint was not achieved: only 1 of 29 patients with paired data met the threshold for functional decline. This paper describes the cohort and biobank as a clinical methods reference for linked translational studies.

### COVID-19 predominance

COVID-19 predominance is the most important contextual feature of this cohort, affecting interpretation of all findings. Sixty-four percent of patients were managed as COVID-19. The cohort is markedly younger (mean age 57 years) and less comorbid than historical hospitalised CAP cohorts, in which mean age typically exceeds 70 years.(19) This reflects the tendency of COVID-19 to affect younger and previously healthier individuals who were more likely to consent and participate. Translational studies using these samples should acknowledge this: immune responses to SARS-CoV-2 are biologically distinct from those to bacterial CAP, and endotype structures identified here may not generalise directly.

The high proportion receiving systemic corticosteroids (64%) reflects dexamethasone use in accordance with COVID-19 management guidelines operative throughout the recruitment period, rather than an unusual approach to bacterial CAP management.

### Phenotyping, endotyping, and subcohort characterisation

The biobank collected here — comprising fresh flow cytometry, transcriptomics, pathogen DNA, and plasma — enables retrospective immune endotyping of this cohort, complementing the clinical phenotyping described above. The COVID vs non-COVID subcohorts within PARIS represent a natural comparison in host response to viral versus bacterial pneumonia, though the small number of non-COVID patients (n=17) limits most formal comparisons. The significantly higher admission CRP observed in the non-COVID group is consistent with the inclusion of bacterial pneumonia, in which CRP is characteristically higher than in viral infection.

The observation that 87% of all enrolled patients met sepsis criteria by 24 hours post enrolment — compared with 68% at the moment of enrolment — illustrates an important point about study design. Using a single-timepoint SOFA score to classify patients, as is common in clinical trials, may misclassify a substantial minority who progress rapidly. This is analogous to the persistence criteria applied in some ARDS trial designs. It also supports the pre-specified decision to combine the two enrolment groups for analysis in this paper.

### Near-absent functional decline

The primary outcome rate of 3.4% (1/29 with paired data) is far below the 20–30% assumed in the sample size calculation.(20) This is almost certainly a true reflection of the population recruited rather than a measurement failure. Pre-pandemic CAP cohorts in which high rates of functional decline are reported are predominantly elderly and multimorbid;(19) a COVID- predominant cohort with mean age 57 years and near-universal pre-morbid independence would be expected to return to baseline activities in the great majority of cases.

The Barthel index measures basic activities of daily living and may lack sensitivity to detect subtler deficits in a younger, higher-functioning population. The reduction in the proportion achieving the maximum DASI score (from 51% to 35%) and the numerically lower median DASI at follow-up suggest that meaningful functional impact was present but below the Barthel detection threshold. A median DASI of 58.2 at enrolment — the maximum possible score — confirms the high pre-morbid functional status of this cohort and the ceiling effect of the Barthel index in this population. Future studies should incorporate more sensitive patient-reported outcome measures alongside traditional indices.(17)

### Persistent systemic inflammation

More than half of survivors with follow-up CRP measurements had persistently elevated levels (>3 mg/L at 6–8 weeks), consistent with published data showing ongoing subclinical inflammation after CAP.(6) The small number of patients with follow-up CRP data (n=16) and the COVID predominance substantially limit the conclusions that can be drawn. Persistent CRP elevation after COVID-19 may reflect distinct processes from those after bacterial pneumonia. No significant association between persistent CRP elevation and persistent radiological opacification was detected, though the numbers were insufficient (n=15 with both variables) for any meaningful inference.

### Mobilisation and nutritional support

Prospective daily data on mobilisation assistance and nutritional support were collected during the inpatient stay. This is an aspect of CAP management that is rarely captured in observational studies, and these data were collected specifically to characterise current practice and inform future interventional trial design. Dietitian input was documented in 91% of patients. Documentation of physiotherapy and mobilisation was more variable, reflecting common challenges in capturing rehabilitation delivery through routine records.

### Strengths and limitations

The principal strengths are the prospective design, the high translational sample collection rates achieved under pandemic pressures (fresh flow cytometry 94%, transcriptomics 97% in the sepsis subgroup, pathogen DNA sampling as detailed in Table 5, plasma 62%), and the detailed clinical characterisation against which biological findings can be interpreted.

Important limitations are as follows. **Sample size and generalisability:** 47 patients at a single centre substantially limits statistical power and external validity. **COVID-19 predominance:** the cohort does not represent the pre-pandemic CAP population for which the study was designed. **Recruitment window:** patients were identified during weekday working hours only, introducing potential selection bias. **Follow-up completeness:** 66% of patients completed follow-up; CRP was available at follow-up in only 34%. **Data quality:** completeness and quality were variable, reflecting the operational pressures of conducting clinical research during a major pandemic. **Primary endpoints not achieved:** neither the PCT metrological objective nor the functional decline endpoint was assessed as planned. **Prospective registration:** the study was not registered on a prospective clinical trial registry. **Medication data:** regular prescribed medications were not comprehensively collected.

### Recommendations for future work

A properly powered multicentre observational study of hospitalised CAP — with adequate follow-up, prospective registration, and robust data collection infrastructure — is needed to test the primary hypotheses that motivated PARIS. Such a study should extend follow-up beyond 6– 8 weeks and incorporate outcome measures sensitive enough to detect impairment in patients who are functionally independent at baseline.

The PARIS biobank provides material for a programme of translational analyses. Future work should aim to identify biological predictors of persistent systemic inflammation and impaired physical recovery following CAP, and to characterise the immunological mechanisms linking acute infection with long-term functional and frailty-related outcomes.

The observation that 44% of patients with available chest radiograph data had persistent opacification at 6–8 weeks warrants further investigation with longer follow-up and linked functional and immunological data, particularly given uncertainty about its prognostic significance following COVID-19 pneumonia.

A better characterisation of the pathophysiological mechanisms linking hospitalisation for CAP with its long-term consequences is a prerequisite for developing targeted interventions — whether pharmacological, rehabilitative, or nutritional — capable of modifying those outcomes.

## Conclusions

The PARIS cohort provides a well-characterised clinical platform and linked translational biobank for studies of pneumonia and sepsis, including immune phenotyping and endotyping. Findings must be interpreted in the context of a pandemic-era, predominantly COVID-19, younger and lower-comorbidity population. Primary protocol endpoints were not achieved. The cohort and biobank underpin a programme of translational immunological studies, for which this paper serves as a clinical methods reference.

## Data Availability

All data produced in the present study are available upon reasonable request to the authors

## Acknowledgements

The authors thank the patients and their families for their participation in this study. We acknowledge the contributions of: Kimberley Lucini and Sinead Donlon (Research Nurses, Royal Surrey NHS Foundation Trust — patient recruitment and sample collection), Dr Ghanem Aldik (Consultant Respiratory Medicine, Royal Surrey NHS Foundation Trust), Dr James Clayton (Consultant Microbiologist, Royal Surrey NHS Foundation Trust), Dr Fernando Martinez Estrada, Dr Natalie Riddell, Dr Shaun McMaster, Kate Penhaligon, and Professor Simon Skene (University of Surrey, statistical advice).

## Artificial Intelligence Assistance

Manuscript drafting was assisted by Claude (Anthropic). All content was verified and approved by the authors, who take full responsibility for accuracy and integrity.

